# Acute temporal effect of ambient air pollution on common congenital cardiovascular defects and cleft palate: a case-crossover study

**DOI:** 10.1101/2022.03.06.22271984

**Authors:** Temuulen Enebish, Meredith Franklin, Rima Habre, Carrie Breton, Nomindelger Tuvshindorj, Gantuya Tumur, Bayalag Munkhuu, David Warburton

## Abstract

This symmetric bidirectional case-crossover study examined the association between short-term ambient air pollution exposure during weeks 3-8 of pregnancy and certain common congenital anomalies in Ulaanbaatar, Mongolia, between 2014 and 2018. Using predictions from a Random Forest regression model, authors assigned daily ambient air pollution exposure of particulate matter <2.5 um aerodynamic diameter, sulphur dioxide, nitrogen dioxide, and carbon monoxide for each subject based on their administrative area of residence. We used conditional logistic regression with adjustment for corresponding apparent temperature to estimate relative odds of select congenital anomalies per IQR increase in mean concentrations and quartiles of air pollutants. The adjusted relative odds of cardiovascular defects (ICD-10 subchapter: Q20-Q28) was 2.64 (95% confidence interval: 1.02-6.87) per interquartile range increase in mean concentrations of particulate matter <2.5 um aerodynamic diameter for gestational week 7. This association was further strengthened for cardiac septal defects (ICD-10 code: Q21, odds ratio: 7.28, 95% confidence interval: 1.6-33.09) and isolated ventricular septal defects (ICD-10 code: Q21.0, odds ratio: 9.87, 95% confidence interval: 1.6-60.93). We also observed an increasing dose-response trend when comparing the lowest quartile of air pollution exposure with higher quartiles on weeks 6 and 7 for Q20-Q28 and Q21 and week 4 for Q21.0. Other notable associations include increased relative odds of cleft lip and cleft palate subchapter (Q35-Q37) and PM_2.5_ (OR: 2.25, 95% CI: 0.62-8.1), SO_2_ (OR: 2.6, 95% CI: 0.61-11.12), and CO (OR: 2.83, 95% CI: 0.92-8.72) in week 4. Our findings contribute to the limited body of evidence regarding the acute effect of ambient air pollution exposure on common adverse birth outcomes.

## Background

Congenital anomalies pose a significant risk of global infant mortality, life-long illness, disability and have a heavy burden on resource-scarce countries. Worldwide estimates show that about 13% of total mortality among children under five years of age was due to congenital anomalies (CA) in 2016 (World Health Organization, 2017). The most common forms of major CA include congenital heart defects, cleft lip and palate and neural tube defects. Although CA are a global problem, 94 percent of births with severe congenital malformations and 95 percent of the mortality among these children occur in middle and low-income countries. Maternal health and other risk factors such as poverty, a higher proportion of older mothers, and a larger frequency of consanguineous marriages partly explain this significant difference. In terms of aetiology, about half of all major CA are unknown and hypothesized to have multiple causal factors, including environmental exposures (Christianson et al., 2005).

Our previous work looking at the short-term effect of air pollution on the occurrence of stillbirth (Enebish et al., 2022) found a significantly increased risk of exposure to high air pollution on days 3 and 6 prior to delivery of a stillbirth. In comparison to stillbirth, there have been a significant number of studies investigating the association between air pollution and CA. Ritz et al. used data from the California Congenital anomalies Monitoring Program to look at associations between CA and average monthly ambient air pollution. To our knowledge, it was the first attempt to explore these associations using case-control design with individual-level data for confounding controls. They found an increased risk of cardiac ventricular septal defects with a dose-response trend as well as increased risks for several other CA (B. R. Ritz et al., 2002). Since then, research groups in the US and around the world have conducted a significant number of case-control studies investigating the association, including Texas (Gilboa et al., 2005), New Jersey (Marshall et al., 2010), California (Padula et al., 2013), Taiwan (Lin et al., 2014), the UK (Dadvand et al., 2011; Dolk et al., 2010; Rankin et al., 2009), Australia (Hansen et al., 2009), Israel (Agay-Shay et al., 2013), Spain (Schembari et al., 2014), and Italy (Gianicolo et al., 2014). The results of these studies varied considerably, probably due to design heterogeneities derived from different case ascertainment procedures and exposure assessment methodologies. A few reviews have attempted to synthesize the findings and found suggestive evidence of increased congenital cardiac anomaly risk associated with NO_2_, SO_2_, and PM_2.5_ (Vrijheid et al., 2011) as well as a significant association between NO_2_ and coarctation of the aorta (Chen et al., 2014). While these studies utilized state or country-wide congenital anomaly surveillance systems for outcome ascertainment, the exposure assessment was usually limited to either spatial gradient based on distance from monitoring stations or temporal gradient based on gestation periods. This kind of exposure assessment may lead to substantial exposure misclassification, exacerbating the residual confounding problem of using registry data with limited covariate information.

Although the biological mechanism of how air pollutants may cause congenital anomalies is unknown, several plausible pathological pathways have been proposed based on laboratory studies. Several studies showed that polycyclic aromatic hydrocarbons (PAH) products attach to DNA, activating DNA repair processes and producing DNA adducts, changing gene function and altering foetal development (Perera et al., 1999). Oxidative stress and inflammatory cytokines triggered by air pollution can adversely affect the foetus via a change in placental functions such as trophoblast proliferation and differentiation as well as apoptosis (Jonakait, 2007; Kannan et al., 2006; Slama et al., 2008). The other possible mechanisms relate to phenotypical changes caused by air pollution due to epigenetic processes such as DNA methylation and acetylation, histone modifications, and microRNA expressions (Baccarelli & Bollati, 2009; Hou et al., 2012). Some studies demonstrated that decreased placental or global methylation is associated with increased PM_2.5_ and PAH (Herbstman et al., 2012; Janssen et al., 2013).

Ulaanbaatar (UB), the capital city of Mongolia, has hazardous levels of air pollution during winter due to more than half of its residents burning coal for domestic heating, combined with an atmospheric inversion that traps emissions within the breathing zone. Infant mortality rates in Mongolia have substantially declined in the last 20 years: 63.4 per 1000 live births in 1990 to 13.4 per 1000 live births in 2018. However, the proportion of deaths due to congenital anomalies has increased from 12.7 percent in 2014 to 16 percent in 2018 (*Comprehensive Survey of the Status of Maternal and Child Morbidity and Mortality, and Prevalence of Congenital Anomalies in Mongolia - V*, 2019). We have seen some alarming results indicating high attributable mortality (Allen et al., 2013) due to air pollution and a strong correlation with miscarriage (Enkhmaa et al., 2014) in the city. Previously, we have mitigated the exposure measurement limitations (inadequate and unreliable monitoring network) of a resource-poor country like Mongolia by utilizing machine learning algorithms for exposure assessment (Enebish et al., 2020). Also, the authors used an electronic surveillance database to ascertain outcomes to overcome the constraints of an overworked and underfinanced healthcare system (Enebish et al., 2022).

The objective of the current study was to investigate the short-term effects of ambient air pollution on the risk of delivering a new-born with select common congenital anomalies in Ulaanbaatar, Mongolia. Specifically, we examined 1) different gestational weeks of exposure periods and 2) risks of select congenital anomalies according to ICD-10 for both continuous and categorical (dose-response) exposure.

## Methods

Ulaanbaatar city registered about 200,000 live births between 2014 and 2018. During this period, there were 1487 cases of CA which resulted in a rate of 7.3 per 1000 live births. This rate is slightly lower than the national average of 8.1 per 1000 live births. The current study includes CA occurring between January 1, 2014, and December 31, 2018, in UB. The Institutional Review Boards at the Health Sciences Campus of USC and the Children’s Hospital Los Angeles and the Medical Ethics Committee of the Ministry of Health of Mongolia approved the study design and methods.

### Study population

We obtained individual-level CA data from the Surveillance Department of the National Center for Maternal and Child Health of Mongolia (NCMCH). In 2014, the Ministry of Health established the department to set up a nationwide active surveillance system for maternal and child health indicators such as maternal mortality, stillbirth, and congenital anomaly. The study team acquired data between 2014 and 2018 from the CA data collection form enumerated in the electronic database. The translated version of the data collection form for CA can be found in Supplemental Materials. The following information was abstracted from the form: maternal age, residential address, education, occupation, parity, smoking status, folic acid usage, date of birth, foetal gender, weight, and gestational age. Gestational age was defined by the best obstetric estimate variable in the birth record, which combines the last menstrual period and ultrasound parameters, as is commonly accepted in clinical practice for gestational age estimation. The WHO defines congenital anomalies as structural or functional anomalies that occur during the intrauterine period and can be identified prenatally, at birth, or sometimes may only be detected later in infancy (Joint WHO-March of Dimes Meeting on Management of Birth Defects and Haemoglobin Disorders (2nd : 2006 : Geneva et al., 2006). For the current study, eligible cases were singleton births, having a maternal residential address in one of the six central districts, and were non-chromosomal CA (Figure 1) having one of 7 CA categories. However, due to a low number of cases, we were only able to analyse five subchapters of ICD-10 chapter Q00-Q99 (Congenital malformations, deformations, and chromosomal abnormalities), one major diagnosis, and one isolated defect (Table 1) in this study (World Health Organization, 2004). We also added Khoroo (small administrative unit of the city) level variables such as percentage of households with internet connectivity (a proxy for socioeconomic status), percentage of Ger (traditional portable dwelling) households, and a number of stoves per square kilometre based on the residential Khoroo of each subject.

**Table 1.** Outcome Groups and ICD-10 Codes Included in the Analysis

| Outcome group | ICD-10 code(s) | n |
| --- | --- | --- |
| Congenital Malformations of The Circulatory System | Q20-Q28 | 282 |
| Congenital Malformations and Deformations of The Musculoskeletal System | Q65-Q79 | 221 |
| Congenital Malformations of Multiple Organ Systems |  | 175 |
| Cleft Lip and Cleft Palate | Q35-Q37 | 159 |
| Congenital Malformations of Eye, Ear, Face and Neck | Q10-Q18 | 131 |
| Congenital malformations of cardiac septa | Q21 | 122 |
| Ventricular septal defect | Q21.0 | 94 |

**Figure 1.**
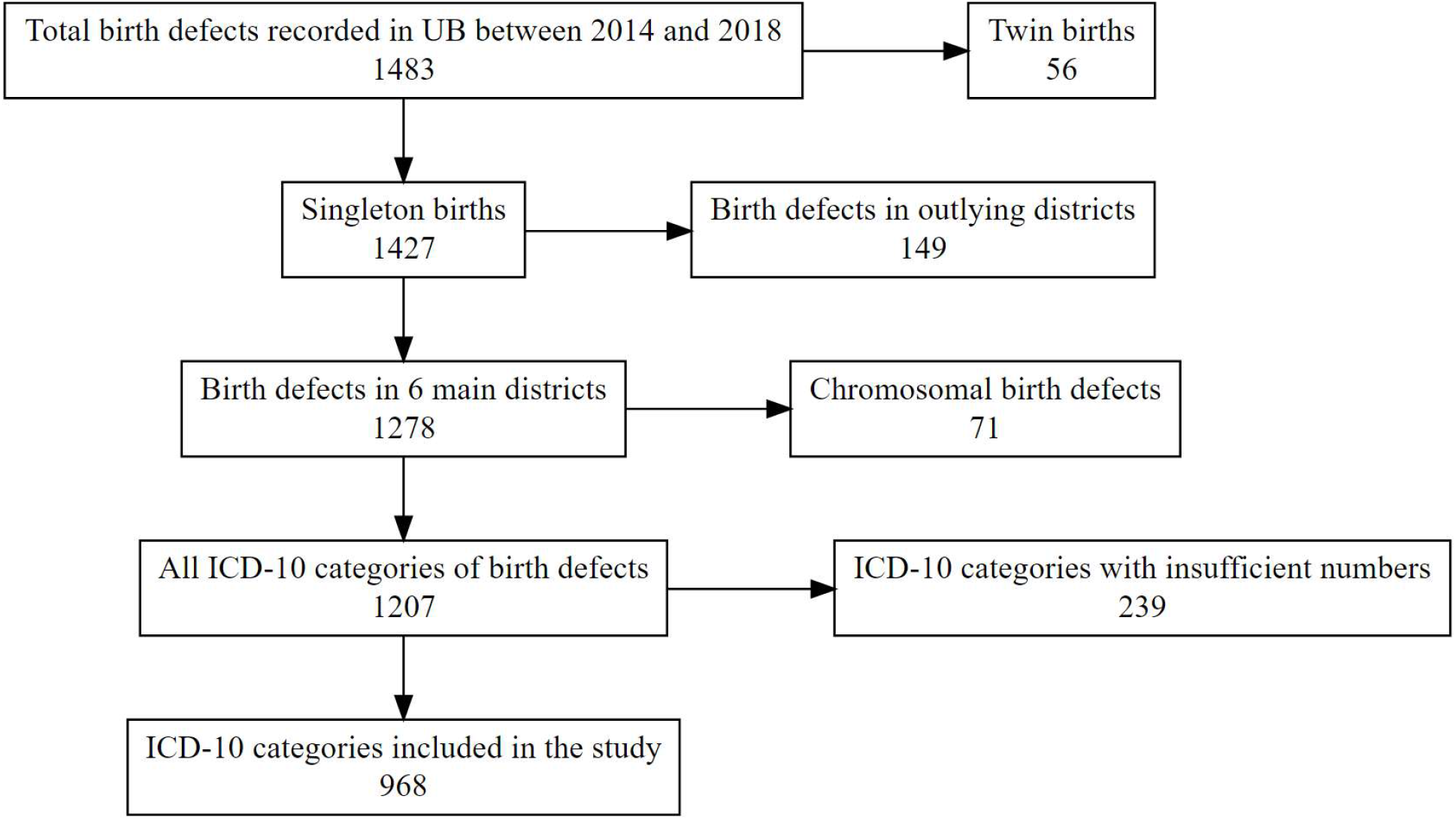
Flow chart of the study population

### Exposure data

We used daily air pollution estimates derived from a previously developed and validated spatiotemporal model, linking them to the residential Khoroo and averaging over the week for the relevant dates of each CA case. We also calculated apparent temperature using atmospheric temperature and wind speed or relative humidity based on the atmospheric temperature of corresponding weeks to control it. The authors covered the details of our prediction models (Enebish et al., 2020) and how we applied them to an adverse reproductive outcome (Enebish et al., 2022) in our previously published works. In addition to continuous exposure measures, we calculated quartiles of each pollutant assigned to our control and case periods to look at the dose-response gradient.

### Study design

The symmetric bidirectional case-crossover design was used to estimate the relative odds of specific CA associated with each interquartile range (IQR) increase in mean pollutant concentrations and 2nd-4th quartile compared to the lowest quartile. We used a case-crossover design with a different referent period selection method in our previous work on the association between stillbirth and ambient air pollution (Enebish et al., 2022). The current study utilizes symmetric bidirectional referent selection instead of time-stratified referent selection. Unlike time-stratified referent selection, bidirectional selection creates non-localizable referent windows, which means that the conditional logistic regression estimating equations are somewhat affected by overlap bias. However, Janes et al. noted that the bias is usually small, particularly for symmetric bidirectional designs (Janes et al., 2005). This control selection method still allows us to control long-term and seasonal time trends by design. We could not use more robust time-stratified referent selection because there is no identifiable single event date for congenital anomalies due to their nature. Instead, we considered susceptible gestational weeks as our case periods and the weekly averages with a one-week wash-out period in both directions as control periods. We looked at gestational weeks 3-8 as susceptible periods since most active foetal organ development happens during this time, thus making them vulnerable to environmental teratogens. For instance, we designated week one and week five as controls when we considered week three as our case period in our analyses. Gestational weeks were determined based on conception dates which we calculated by subtracting gestational weeks at birth from the birth dates of each subject.

### Statistical analysis

We used conditional logistic regression to estimate the effect of ambient air pollution (PM_2.5_, SO_2_, NO_2_, and CO) on the occurrence of the selected CA during vulnerable periods of gestation. Like our previous study on stillbirth, we estimated log odds of each CA per IRQ increase in continuous measures and quartiles with the lowest as a reference in categorical measures. All models were adjusted for the weekly mean apparent temperature of corresponding gestational weeks to control time-dependent confounding. Confounders such as age, socioeconomic status, and comorbidity were considered unchanged in the short term and therefore controlled by study design. Ambient air pollution exposure quartiles were examined to determine whether there are identifiable dose-response curves for any of the CA and determine their shape. We evaluated influential gestational weeks of exposure based on the magnitude and pattern of the observed risk estimates and their confidence interval widths. All statistical modelling and geospatial computing were performed in R version 3.6.2 (R Core Team, 2019) using packages “tidyverse” (Wickham et al., 2019), “sf” (Pebesma, 2018), and “survival” (Therneau & Grambsch, 2000).

## Results

After applying the inclusion and exclusion criteria, our study population consisted of 968 congenital anomaly cases between 2014 and 2018 (Figure 1). We show the demographic and residential factors of the study subjects by CA groups in Table 2. About one in five mothers were 35 years or older, and most of them had graduated college or trade school (61%) and were employed (63%). Only about 6% of the mothers reported smoking, while 43% had used folic acid supplements within the first 12 weeks of gestation. Most mothers had previously given birth (65%), and the foetal gender was majority male (58%). Regarding residential Khoroo characteristics, more mothers were from Khoroos with less connectivity to the internet, more Ger households, and higher stove density. There were no substantial differences between CA groups for any demographic or residential characteristics.

**Table 2.**
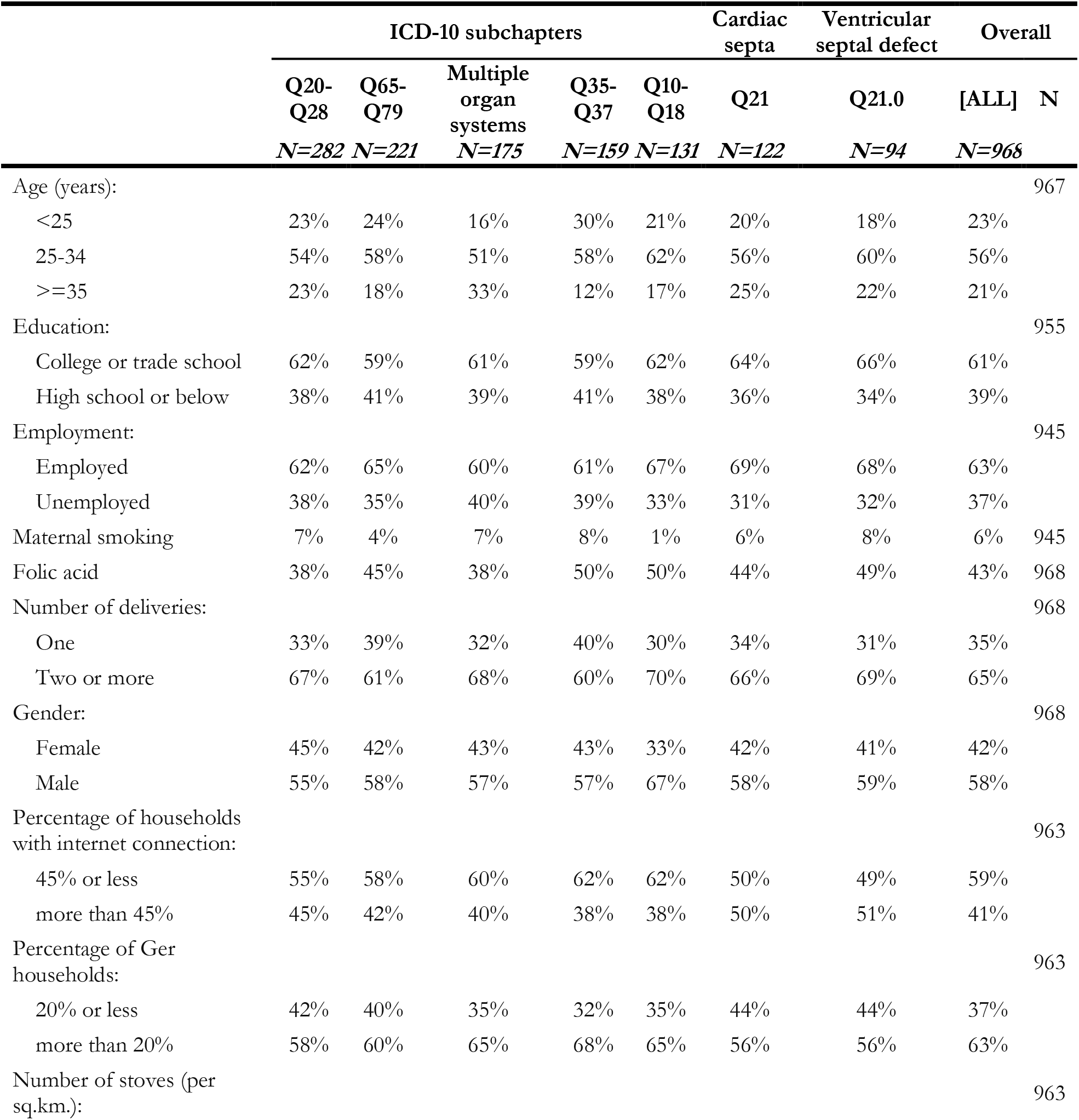
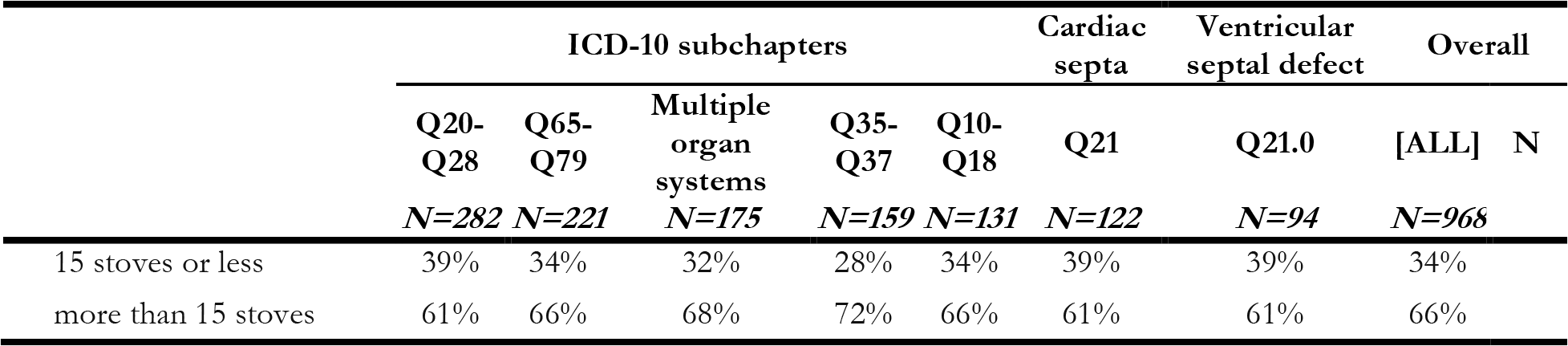
Distribution of Covariates Among Pooled Congenital Anomaly Cases, and Select Outcome Groups, Ulaanbaatar, 2014–2018

Odds ratio (OR) estimates and their 95% confidence intervals (95% CI) for each IQR increase in mean pollutant concentrations for every CA group examined by different gestational weeks are shown in Tables 3 and 4. We observed significantly increased relative odds (OR: 2.64, 95% CI: 1.02-6.87) of cardiovascular defects (Q20-Q28) per IQR increase in mean PM_2.5_ concentration in week 7 (Table 3). This association with PM_2.5_ was strengthened in the subgroup of cardiac septal defects (OR: 7.28, 95% CI: 1.6-33.09) and isolated ventricular septal defects (OR: 9.87, 95% CI: 1.6-60.93). We also obtained similar significantly increased relative odds of cardiac septal defects (OR: 3.12, 95% CI: 0.99-9.82) and isolated ventricular septal defects (OR: 3.96, 95% CI: 1.05-14.93) per IQR increase in mean concentrations of CO in week 7. The relative odds of the association between PM_2.5_ and ventricular septal defects were increased on week 4 (OR: 6.01, 95% CI: 1.04-34.7) as well (Table 3). Other notable associations include increased relative odds of cleft lip and cleft palate subchapter (Q35-Q37) and PM_2.5_ (OR: 2.25, 95% CI: 0.62-8.1), SO_2_ (OR: 2.6, 95% CI: 0.61-11.12), and CO (OR: 2.83, 95% CI: 0.92-8.72) in week 4 (Table 4). We did not see consistent effects on any of the other ICD-10 CA subchapters.

**Table 3.**
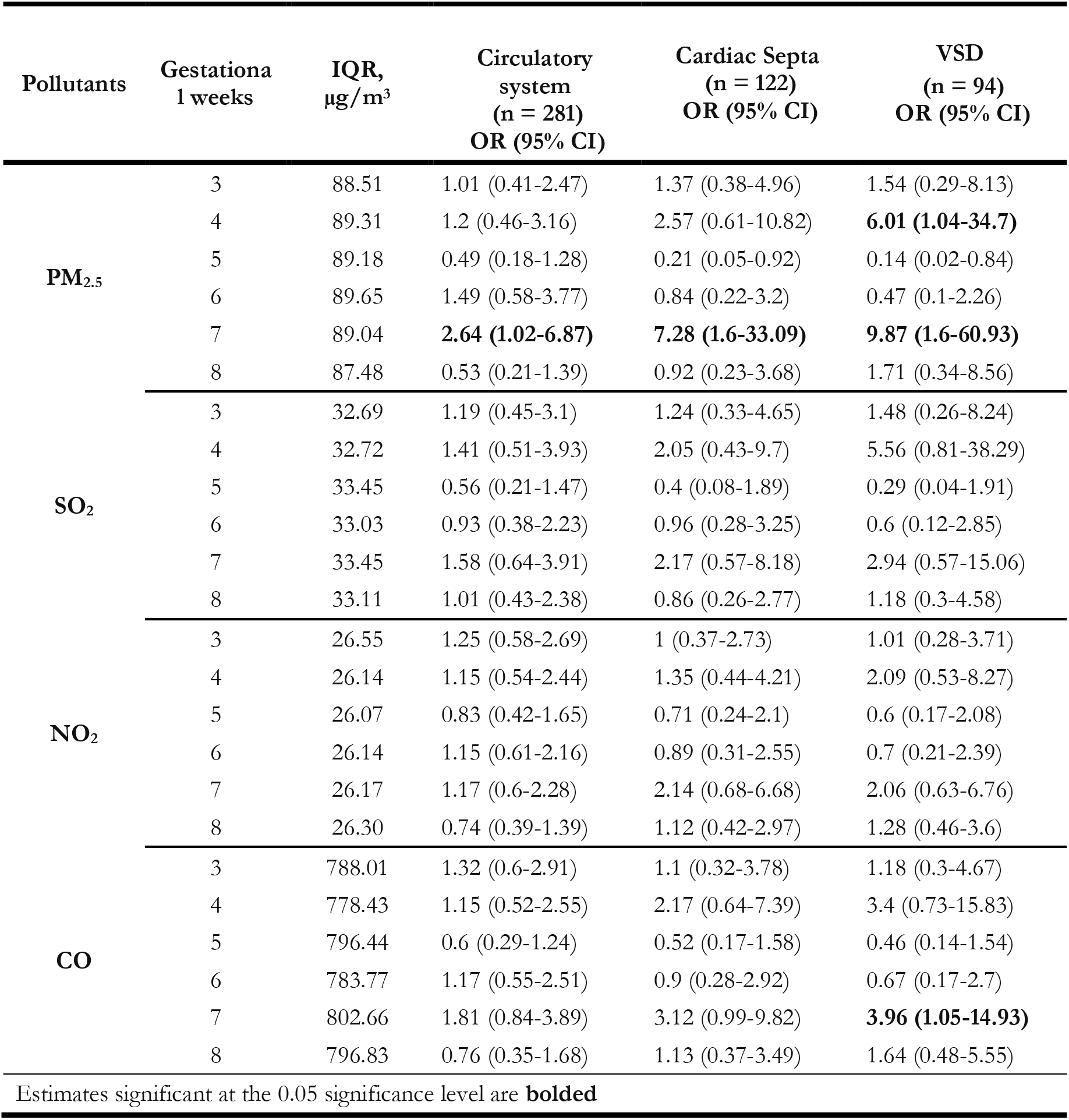
Relative Odds of Congenital Anomalies Related to Cardiovascular System

**Table 4.**
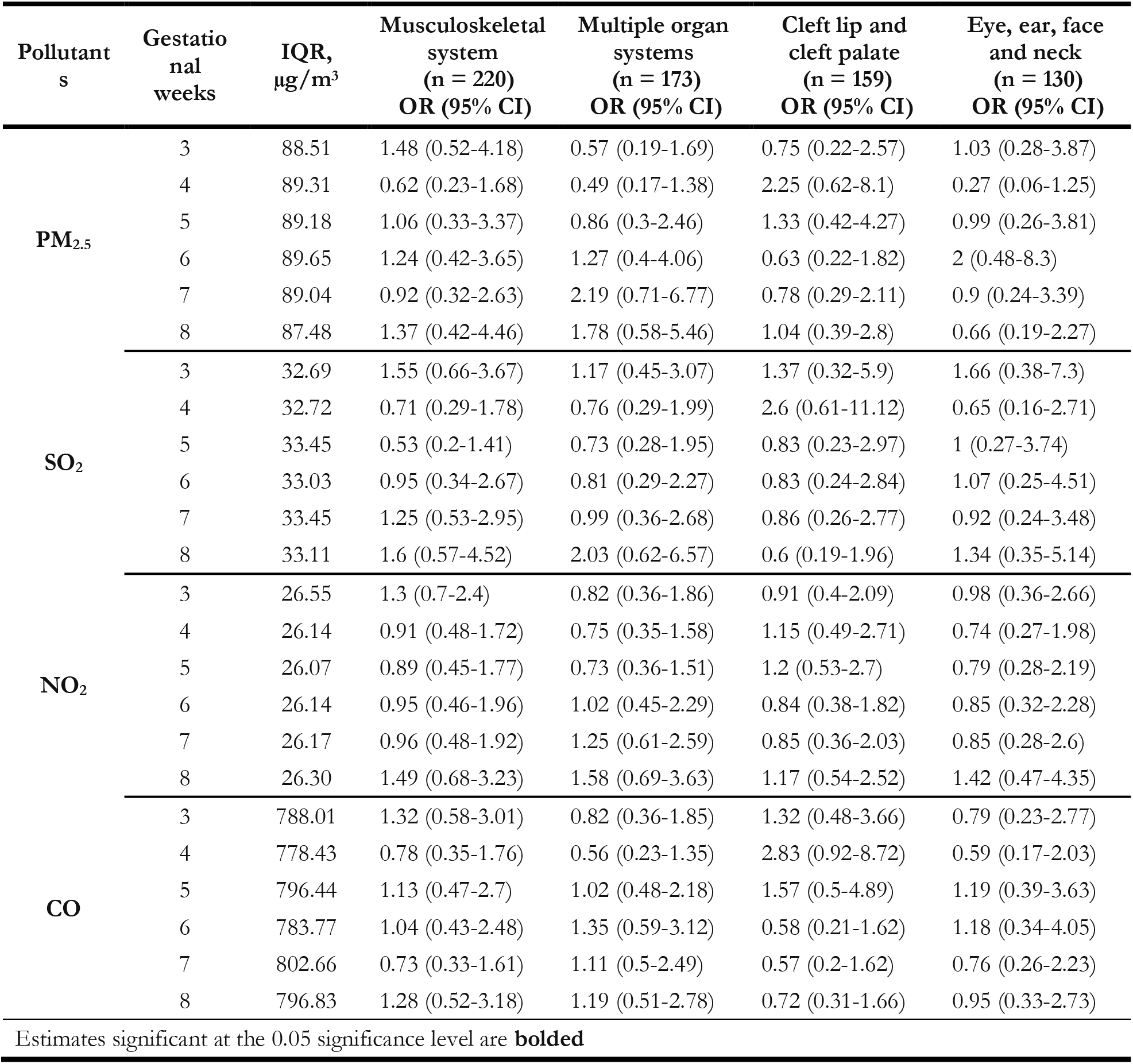
Relative Odds of Select Congenital Anomalies

| Pollutants | Gestational weeks | IQR, $\mu\text{g}/\text{m}^3$ | Musculoskeletal system (n = 220)<br>OR (95% CI) | Multiple organ systems (n = 173)<br>OR (95% CI) | Cleft lip and cleft palate (n = 159)<br>OR (95% CI) | Eye, ear, face and neck (n = 130)<br>OR (95% CI) |
| --- | --- | --- | --- | --- | --- | --- |
| PM<sub>2.5</sub> | 3 | 88.51 | 1.48 (0.52-4.18) | 0.57 (0.19-1.69) | 0.75 (0.22-2.57) | 1.03 (0.28-3.87) |
|  | 4 | 89.31 | 0.62 (0.23-1.68) | 0.49 (0.17-1.38) | 2.25 (0.62-8.1) | 0.27 (0.06-1.25) |
|  | 5 | 89.18 | 1.06 (0.33-3.37) | 0.86 (0.3-2.46) | 1.33 (0.42-4.27) | 0.99 (0.26-3.81) |
|  | 6 | 89.65 | 1.24 (0.42-3.65) | 1.27 (0.4-4.06) | 0.63 (0.22-1.82) | 2 (0.48-8.3) |
|  | 7 | 89.04 | 0.92 (0.32-2.63) | 2.19 (0.71-6.77) | 0.78 (0.29-2.11) | 0.9 (0.24-3.39) |
|  | 8 | 87.48 | 1.37 (0.42-4.46) | 1.78 (0.58-5.46) | 1.04 (0.39-2.8) | 0.66 (0.19-2.27) |
| SO<sub>2</sub> | 3 | 32.69 | 1.55 (0.66-3.67) | 1.17 (0.45-3.07) | 1.37 (0.32-5.9) | 1.66 (0.38-7.3) |
|  | 4 | 32.72 | 0.71 (0.29-1.78) | 0.76 (0.29-1.99) | 2.6 (0.61-11.12) | 0.65 (0.16-2.71) |
|  | 5 | 33.45 | 0.53 (0.2-1.41) | 0.73 (0.28-1.95) | 0.83 (0.23-2.97) | 1 (0.27-3.74) |
|  | 6 | 33.03 | 0.95 (0.34-2.67) | 0.81 (0.29-2.27) | 0.83 (0.24-2.84) | 1.07 (0.25-4.51) |
|  | 7 | 33.45 | 1.25 (0.53-2.95) | 0.99 (0.36-2.68) | 0.86 (0.26-2.77) | 0.92 (0.24-3.48) |
|  | 8 | 33.11 | 1.6 (0.57-4.52) | 2.03 (0.62-6.57) | 0.6 (0.19-1.96) | 1.34 (0.35-5.14) |
| NO<sub>2</sub> | 3 | 26.55 | 1.3 (0.7-2.4) | 0.82 (0.36-1.86) | 0.91 (0.4-2.09) | 0.98 (0.36-2.66) |
|  | 4 | 26.14 | 0.91 (0.48-1.72) | 0.75 (0.35-1.58) | 1.15 (0.49-2.71) | 0.74 (0.27-1.98) |
|  | 5 | 26.07 | 0.89 (0.45-1.77) | 0.73 (0.36-1.51) | 1.2 (0.53-2.7) | 0.79 (0.28-2.19) |
|  | 6 | 26.14 | 0.95 (0.46-1.96) | 1.02 (0.45-2.29) | 0.84 (0.38-1.82) | 0.85 (0.32-2.28) |
|  | 7 | 26.17 | 0.96 (0.48-1.92) | 1.25 (0.61-2.59) | 0.85 (0.36-2.03) | 0.85 (0.28-2.6) |
|  | 8 | 26.30 | 1.49 (0.68-3.23) | 1.58 (0.69-3.63) | 1.17 (0.54-2.52) | 1.42 (0.47-4.35) |
| CO | 3 | 788.01 | 1.32 (0.58-3.01) | 0.82 (0.36-1.85) | 1.32 (0.48-3.66) | 0.79 (0.23-2.77) |
|  | 4 | 778.43 | 0.78 (0.35-1.76) | 0.56 (0.23-1.35) | 2.83 (0.92-8.72) | 0.59 (0.17-2.03) |
|  | 5 | 796.44 | 1.13 (0.47-2.7) | 1.02 (0.48-2.18) | 1.57 (0.5-4.89) | 1.19 (0.39-3.63) |
|  | 6 | 783.77 | 1.04 (0.43-2.48) | 1.35 (0.59-3.12) | 0.58 (0.21-1.62) | 1.18 (0.34-4.05) |
|  | 7 | 802.66 | 0.73 (0.33-1.61) | 1.11 (0.5-2.49) | 0.57 (0.2-1.62) | 0.76 (0.26-2.23) |
|  | 8 | 796.83 | 1.28 (0.52-3.18) | 1.19 (0.51-2.78) | 0.72 (0.31-1.66) | 0.95 (0.33-2.73) |

Estimates significant at the 0.05 significance level are **bolded**

Estimates of relative odds and their 95% CI derived from comparing the lowest quartile of PM_2.5_ concentration with 2nd-4th quartiles are presented in Figures 2-6. Our primary goal was to visually show the general trend of dose-response for different types of CA instead of specific numbers due to the instability of effect estimates caused by a small sample size. For brevity, we show estimates of the short-term effect of PM_2.5_ on cardiovascular, musculoskeletal, cardiac septal, and ventricular septal defects (VSD) as examples. We observed dose-response patterns for weeks 6 and 7 for cardiovascular and cardiac septal defects, but for VSD, the increasing dose-response trend occurred in week 4. A less pronounced dose-response trend was observed in weeks 4 and 8 for musculoskeletal anomalies and cleft lip and palate. We found similarly suggestive dose-response patterns in different pollutants, gestational weeks, and congenital anomalies (results not shown).

**Figure 2.**
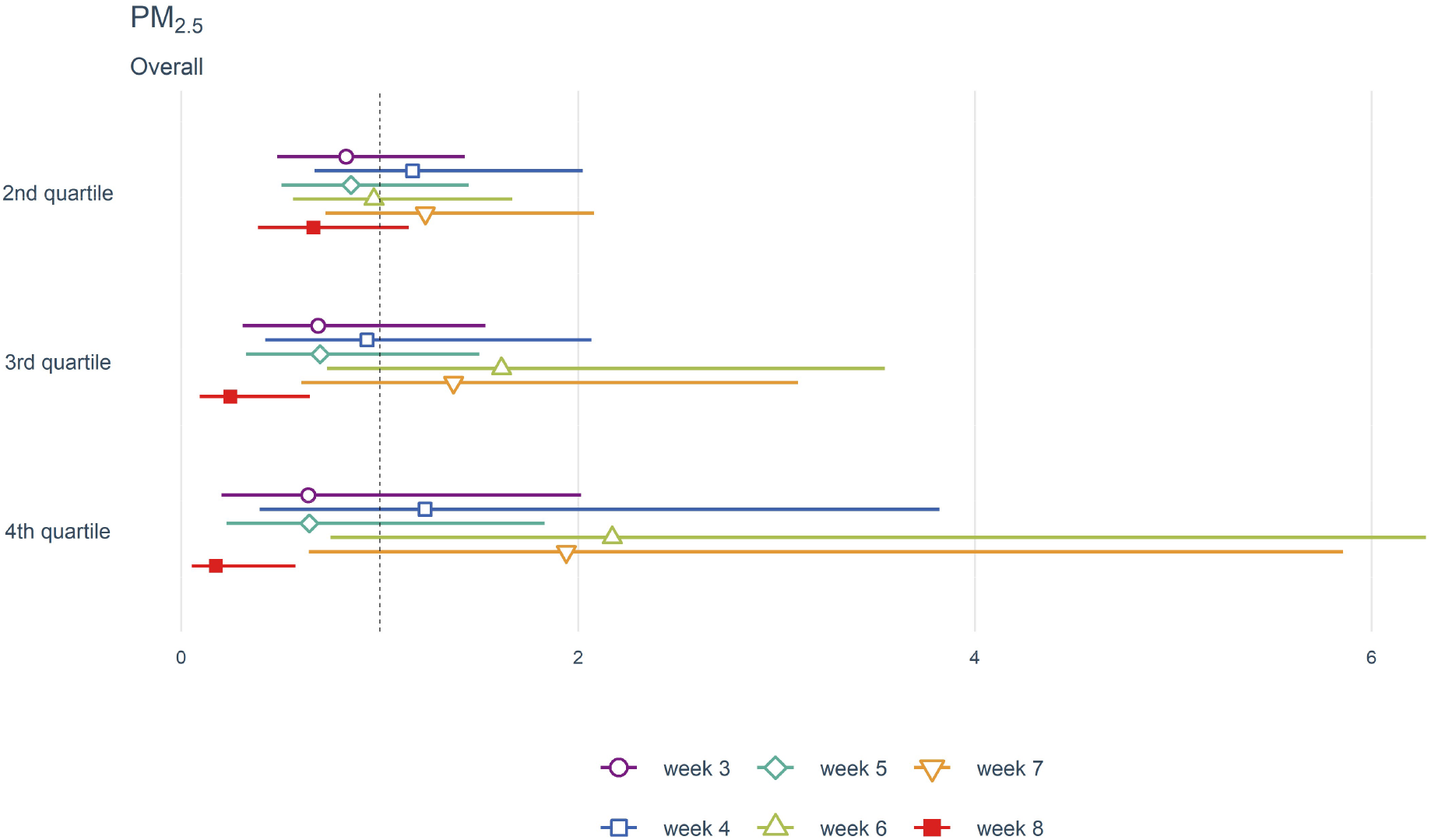
Effect estimates of air pollution on cardiovascular anomaly risk

## Discussion

We found a considerably increased short-term risk of cardiovascular defects, particularly cardiac septal defects, associated with per IQR increase in mean concentrations of PM_2.5_ and CO in gestational week 7. This association was further observed to have dose-response gradients when using categorical exposure measures. Although there have been multiple studies looking at the association between ambient air pollution and congenital anomalies in recent years (Girguis et al., 2016; Huang et al., 2019; Lavigne et al., 2019; Vinceti et al., 2016; Zhang et al., 2016; Zhao et al., 2018), our results contribute substantially to the evidence base on short-term effects of air pollution on congenital anomalies by looking at susceptible periods of foetal development at fine temporal detail. To our knowledge, this is the first study to utilize case-crossover design to control for time-invariant confounders in examining the association between ambient air pollution and congenital anomalies.

The findings of our study suggest that there may be differential effects of air pollutants within the critical period of foetus development (weeks 3-8). This is corroborated by embryological studies on lab animals showing specific stages of organ development. For instance, neural crest cells start migrating to develop endocardial tubes at the start of the fourth week, resulting in ventricular septation and outflow tract formation by weeks 7 and 8 in cardiac development (Gittenberger-de Groot et al., 2005). There is also evidence of teratogen-induced oxidative stress in earlier weeks affecting later developments of cardiac structures via apoptosis among migrating cells (Morgan et al., 2008) or different pathway of neural crest cells (Jain et al., 2011). Therefore, it is possible that harmful air pollutant exposure at a particular gestational week may not present itself in the form of CA until later weeks. Additional experimental studies to elucidate this relationship are needed.

In contrast to previous studies, we could not look at isolated congenital anomalies except for VSD due to the small sample size in each of the ICD-10 subchapters. This is important since it is unlikely that a specific air pollutant would affect all types of anomalies and that even single, isolated anomalies could have multifactorial aetiology (B. Ritz, 2010). With that said, our finding of an association between cardiovascular defects and particulate matter is consistent with several earlier case-control studies (Agay-Shay et al., 2013; Huang et al., 2019; Padula et al., 2013; Stingone et al., 2014; Zhang et al., 2016). Most of the previous literature on the topic considered somewhat coarse temporal resolution when it comes to vulnerable periods of development by using either monthly averages of the first three months of pregnancy or average of gestational weeks 3-8. Below we compare our results to a few studies examining specific gestational week exposures. Stingone et al. examined maternal exposure to criteria air pollutants and congenital heart defects using the US National Congenital Anomalies Prevention Study data. They used both 1-week and 7-week averages of pollutants using the closest monitoring station to the maternal residence and found several individual exposure-weeks associations that were not identified from 7-week averages (Stingone et al., 2014). Specifically, the association between VSD and particulate matter was stronger in week three than in week 4 in our categorical exposure findings (Figure 6). Two studies from Wuhan, China, looked at the association between air pollution and the risk of congenital heart defects (Zhang et al., 2016) and oral clefts (Zhao et al., 2018). Zhang et al. found monotonically increasing relative odds of VSD per 10*μ*g/m^3^ increase in mean PM_2.5_ concentration during gestational weeks 7-10. This is somewhat comparable to our finding of significant association between VSD and per IQR increase in the mean concentration of PM_2.5_ in week 7. On the other hand, Zhao et al. reported an increased risk of oral cleft associated with per 10*μ*g/m^3^ change in PM_2.5_ in gestational weeks 4-9. In contrast, we found increased relative odds of cleft lip and cleft palate subchapter and PM_2.5_, SO_2_, and CO in week 4 (Table 4).

**Figure 3.**
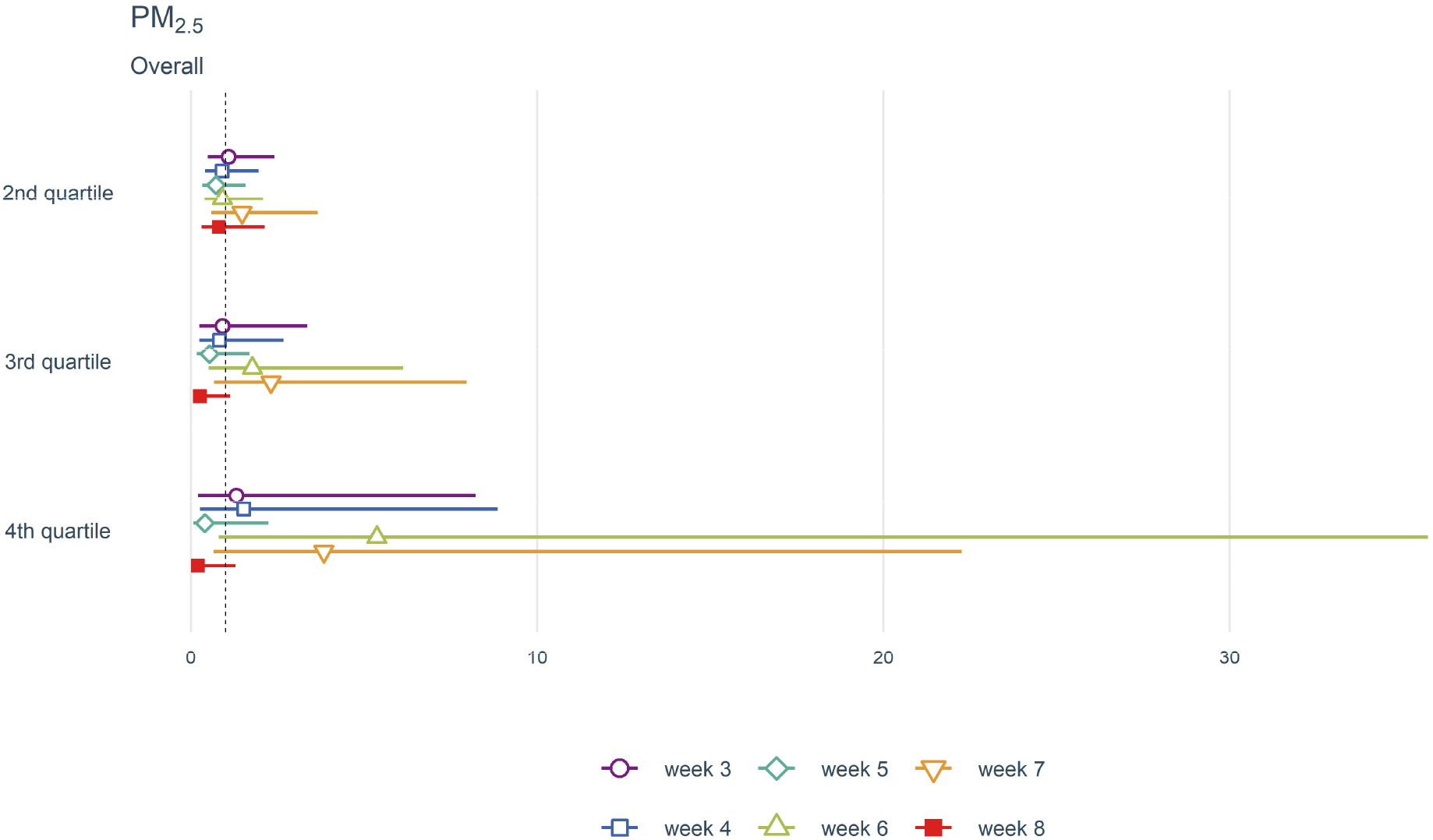
Effect estimates of air pollution on cardiac septal anomaly risk

**Figure 4.**
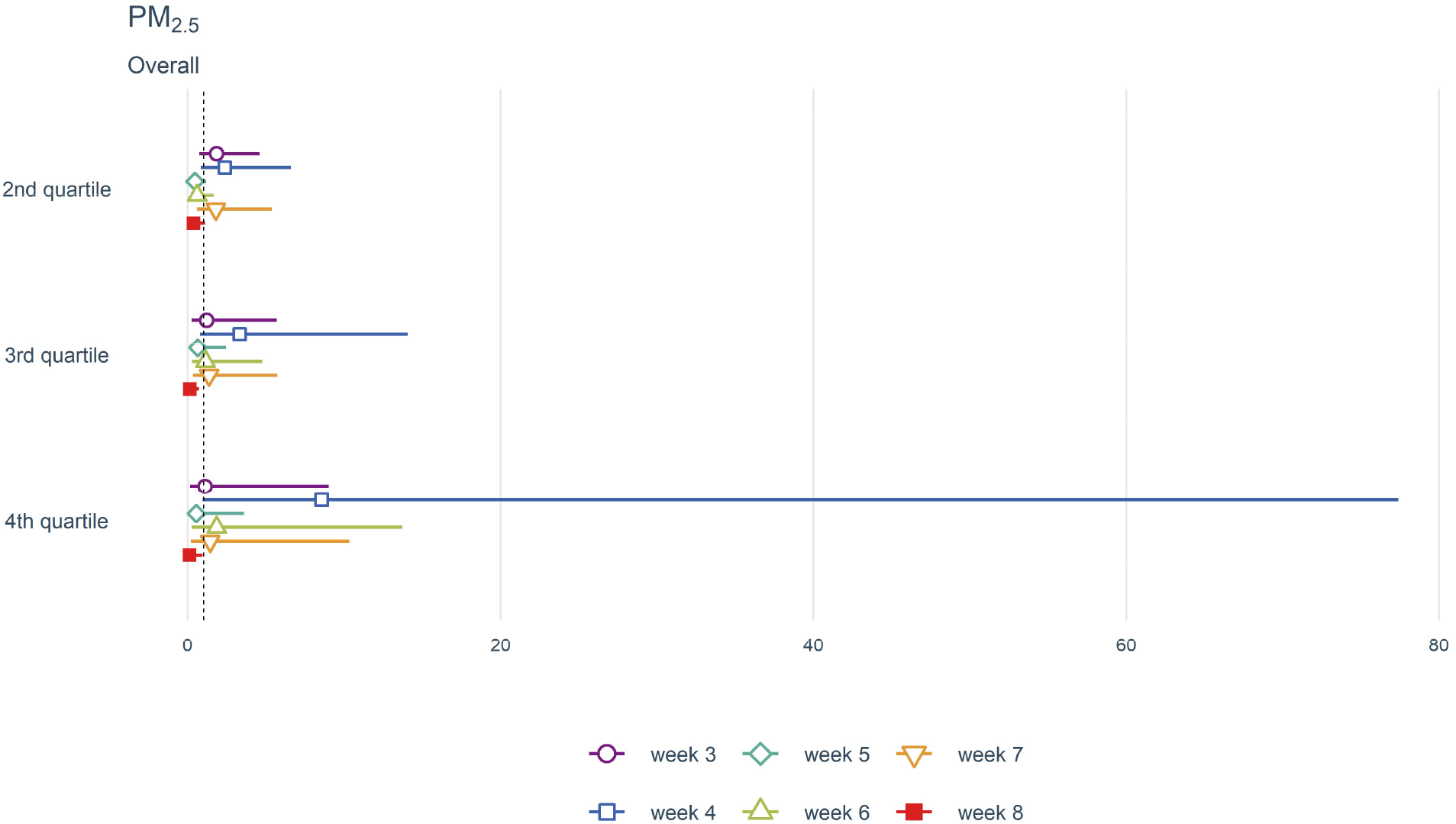
Effect estimates of air pollution on ventricular septal defect risk

**Figure 5.**
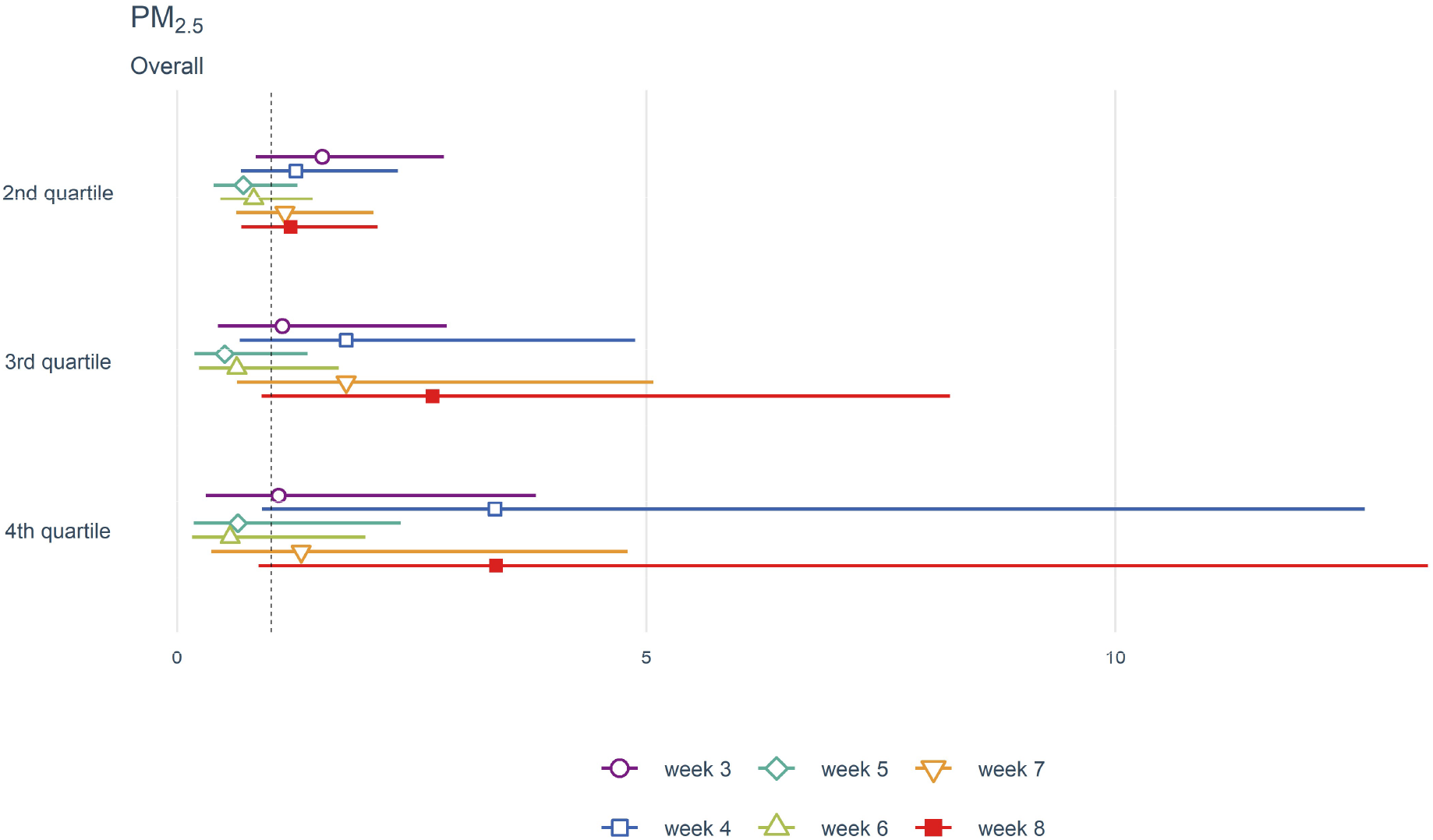
Effect estimates of air pollution on musculoskeletal anomaly risk

**Figure 6.**
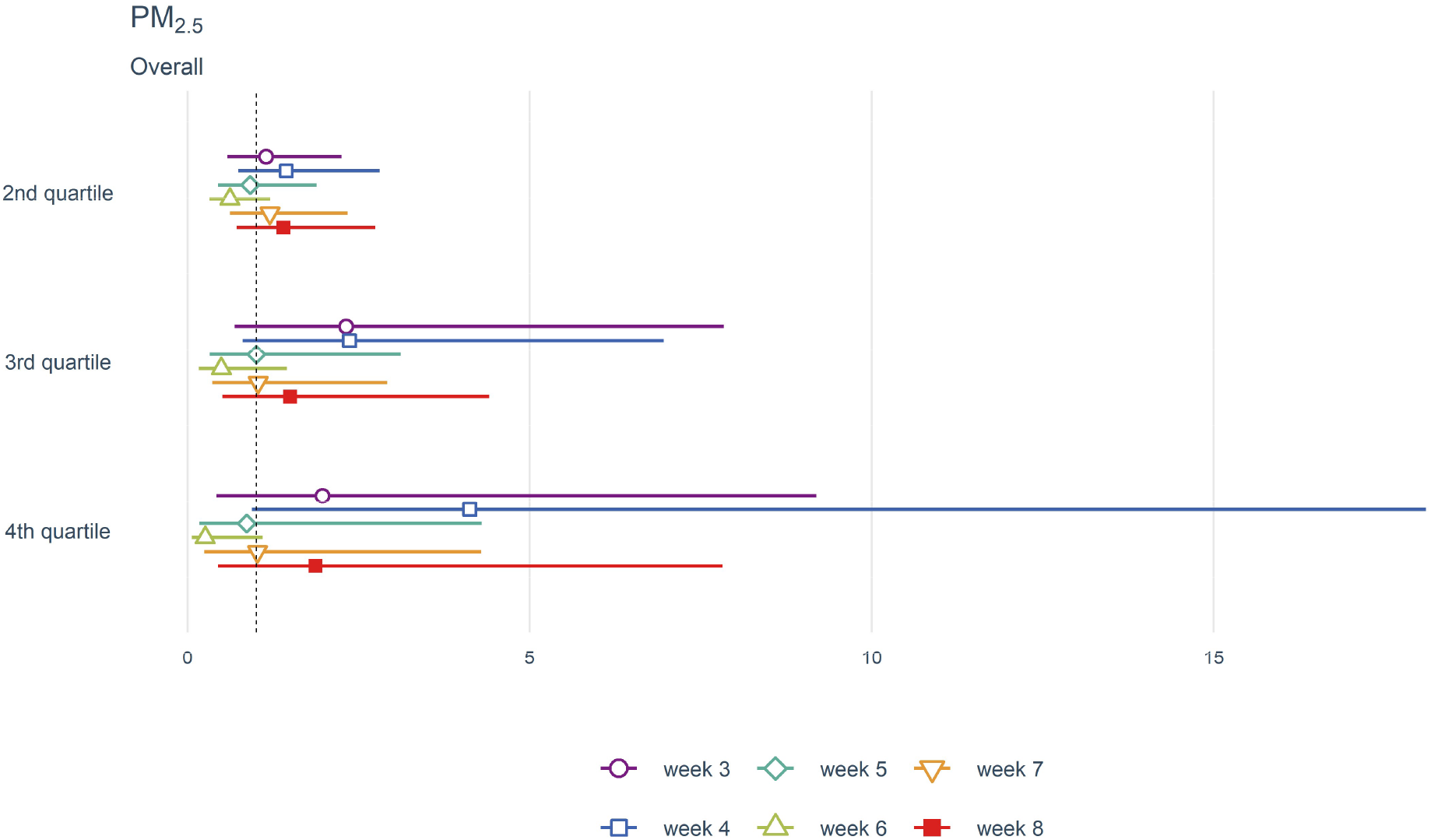
Effect estimates of air pollution on cleft lip and cleft palate risk

In order to interpret our findings in the context of current literature, we must keep in mind the emission source of air pollution in UB. As we noted before, the dominant source is domestic coal combustion due to the need for heating and cooking in Ger districts during harsh winters exacerbated by atmospheric inversion and lower mixing height during cold seasons. This is an important and distinct emissions source as compared to manufacturing in developing countries and vehicular emission in developed countries. The composition of particulate matter may be substantially different from compositions in other countries, and this may differentially affect the susceptible windows of organ development with regards to specific CA. Considering this and the relatively homogenous population of Mongolia, the generalizability of our study results may be limited. However, despite these differences, it is suggestive to see partially consistent results with previous studies.

Estimating personal exposure via prediction modelling based on scarce ground-level monitors will inevitably lead to measurement error in exposure regardless of the model performance. In addition, we were only able to predict air pollution exposure based on the residential administrative unit (Khoroo) at the time of delivery. However, a recent study on the effect of residential mobility during pregnancy on air pollution exposure misclassification found a minor impact (Warren et al., 2018). Measurement errors resulting from the above reasons will be non-differential and will likely lead to underestimating the risk. The other limitation of our study was the inadequate sample size for investigating the effect of air pollutants on isolated congenital anomalies rather than ICD-10 subchapters that covered an extensive range of congenital anomalies with possibly differing aetiologies. This was unavoidable due to the relatively recent establishment of the National Surveillance Department in Mongolia and will only strengthen as time goes by.

This study observed increased relative odds of select CA with short-term, higher exposure to air pollutants at specific periods of gestation critical for organogenesis. Some of the associations were only on specific weeks. They had a dose-response trend, which suggests that accounting for temporal and spatial variability may lead to a better understanding of how and when air pollutants affect CA risk. Future studies in this research area should focus on improving exposure assessment, incorporating a multipollutant approach, and ascertaining more isolated congenital anomalies to facilitate the investigation into different susceptible windows of various congenital anomalies.

## Data Availability

All data produced in the present study are available upon reasonable request to the authors.

